# A prospective evaluation of three saliva qualitative antigen testing kits for the detection of SARS-CoV-2 in Japan

**DOI:** 10.1101/2022.12.18.22281291

**Authors:** Norihiko Terada, Yusaku Akashi, Yuto Takeuchi, Atsuo Ueda, Shigeyuki Notake, Koji Nakamura, Hiromichi Suzuki

**Affiliations:** Department of Infectious Diseases, Faculty of Medicine, University of Tsukuba, 1-1-1 Tennodai, Tsukuba, Ibaraki 305-8575, Japan; Division of Infectious Diseases, Department of Medicine, Tsukuba Medical Center Hospital, 1-3-1 Amakubo Tsukuba, Ibaraki 305-8558, Japan; Akashi Internal Medicine Clinic, 3-1-63 Asahigaoka, Kashiwara, Osaka 582-0026, Japan; Department of Infectious Diseases, University of Tsukuba Hospital, 2-1-1 Amakubo, Tsukuba, Ibaraki 305-8576, Japan; Department of Clinical Laboratory, Tsukuba Medical Center Hospital, 1-3-1 Amakubo, Tsukuba, Ibaraki 305-8558, Japan

**Keywords:** Saliva sample, SARS-CoV-2, COVID-19, qualitative antigen testing

## Abstract

**Introduction:** Rapid qualitative antigen testing has been widely used for the laboratory diagnosis of COVID-19 with nasopharyngeal samples. Saliva samples have been used as alternative samples, but the analytical performance of those samples for qualitative antigen testing has not been sufficiently evaluated.

**Methods:** A prospective observational study evaluated the analytical performance of three In Vitro Diagnostics (IVD) approved COVID-19 rapid antigen detection kits for saliva between June 2022 and July 2022 in Japan using real-time reverse transcription polymerase chain reaction (RT-PCR) as a reference. A nasopharyngeal sample and a saliva sample were simultaneously obtained, and RT-PCR was performed.

**Results:** In total, saliva samples and nasopharyngeal samples were collected from 471 participants (140 RT-PCR-positive saliva samples and 143 RT-PCR-positive nasopharyngeal samples) for the analysis. The median Ct values were 25.5 (interquartile range [IQR]: 21.9-28.8) for saliva samples and 17.1 (IQR: 15.5-18.7) for nasopharyngeal samples (p<0.001). Compared with saliva samples of RT-PCR, the sensitivity and specificity were 46.4% and 99.7% for ImunoAce SARS-CoV-2 Saliva, 59.3% and 99.1% for Espline SARS-CoV-2 N, and 61.4% and 98.8% for QuickChaser Auto SARS-CoV-2, respectively. The sensitivity is >90% for saliva samples with a moderate-to-high viral load (Ct<25), whereas the sensitivity is <70% for high-viral-load nasopharyngeal samples (Ct<20).

**Conclusion:** COVID-19 rapid antigen detection kits with saliva showed high specificities, but the sensitivities varied among kits, and the analytical performance of saliva qualitative antigen detection kits was much worse than that of kits using nasopharyngeal samples.

## Introduction

The proper diagnosis of COVID-19 is critical for infection control, and the gold-standard test for such a diagnosis is a nucleic acid amplification test with reverse transcription polymerase chain reaction (RT-PCR) using nasopharyngeal (NP) specimens [1]. However, it can take hours to receive results from RT-PCR after sample submission, and the specimen collection procedure of NP samples requires special handling by healthcare professionals and induces significant discomfort in the patient as well as coughing and sneezing [2]. This limits its application in household and community settings [3].

Qualitative antigen tests, which have an easy-to-perform specimen-handling procedure, wide availability, short performance time, have been developed as an alternative to RT-PCR [4], and NP samples and anterior nasal samples have been used for testing in both symptomatic and asymptomatic patients [5]. Saliva samples have also been widely used as samples for RT-PCR and quantitative antigen tests [6], but their diagnostic performance has been considered insufficient for qualitative antigen tests [7].

In 2022, several saliva qualitative antigen detection kits were newly developed, and In Vitro Diagnostics (IVD) approval was given in Japan. However, their analytical performance was evaluated only by the manufacturers. We therefore conducted prospective evaluations of three IVD-approved saliva antigen qualitative testing kits.

## Method

This study was performed with samples submitted by both symptomatic and asymptomatic patients between June 8, 2021, and July 12, 2022, at a drive-through PCR center at Tsukuba Medical Center Hospital (TMCH), which intensively performed COVID-19 PCR evaluations with NP samples or saliva samples in the Tsukuba district of Tsukuba, Ibaraki Prefecture, Japan. People with and without symptoms were referred from 49 clinics and a local public health center during the study period. Asymptomatic individuals had a history of contact with confirmed or suspected COVID-19 cases.

All evaluations were performed after informed consent was obtained. The informed consent process was performed verbally with documentation in the patient’s electronic medical record in order to prevent infection transmission. The ethics board of TMCH (approval number:2021-055) approved the protocol.

### Study process

NP and saliva samples were simultaneously obtained from participants. The sample collection was performed as previously described [2,8-19]. All antigen tests for saliva were immediately performed on site after sample collection. Each antigen test was performed based on the manufacturer’s instructions in the package insert (Fig. 1). In cases with a poor control response, a re-test was performed. After the antigen evaluation, all saliva samples were preserved at −80 °C until reference RT-PCR.

**Figure 1.**
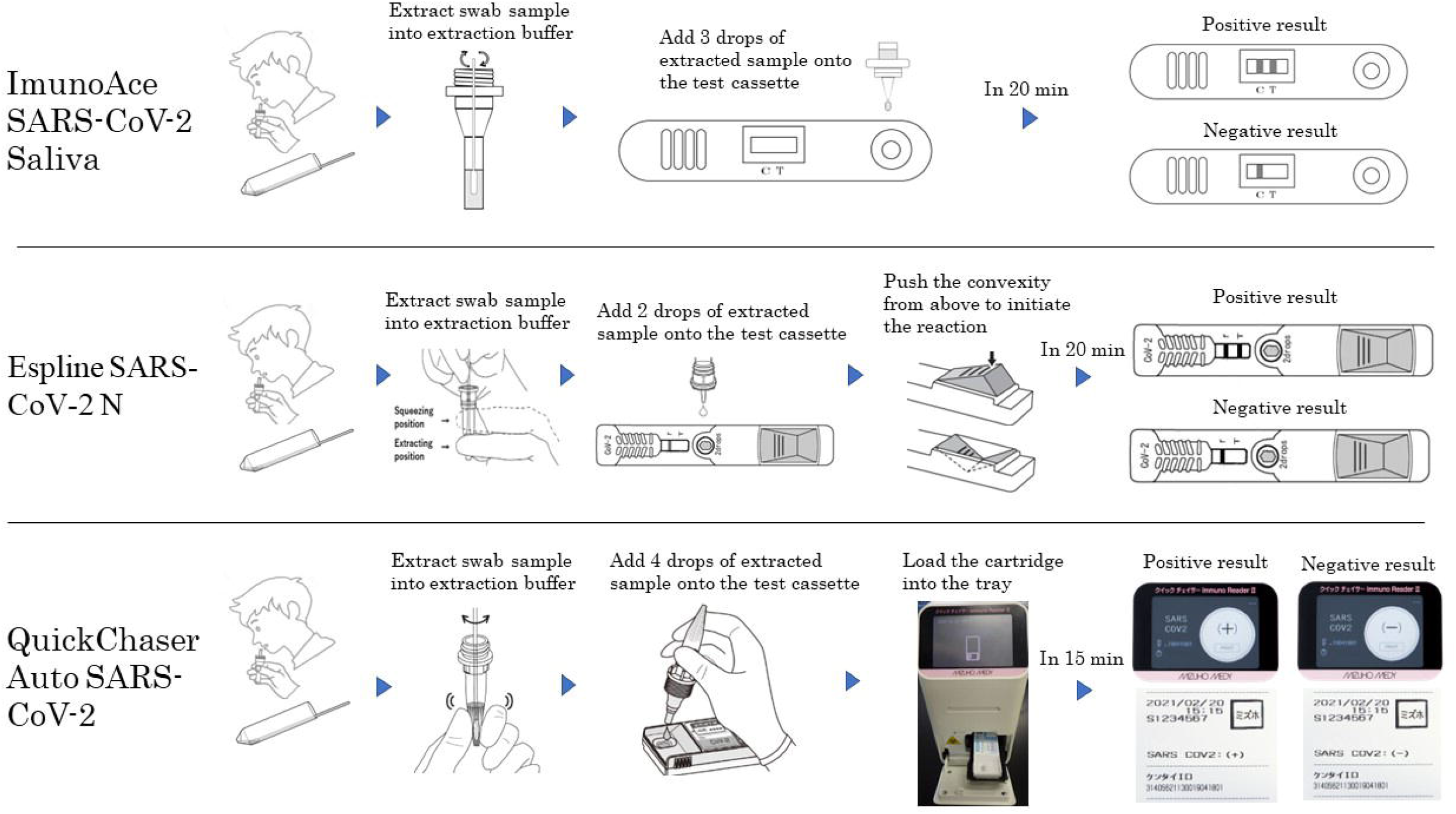
Test flow diagram of each rapid qualitative antigen testing kit for the detection of SARS-CoV-2 in saliva. The photo and illustration of Espline SARS-CoV-2 N were provided by Fujirebio Holdings, Inc. The photo and illustration of QuickChaser Auto SARS-CoV-2 were provided by MIZUHO MEDY Co., Ltd.

Each NP swab was diluted in 3 mL of Universal Transport Medium (Copan Italia S.p.A., Brescia, Italy) on site, and the sample was transferred to the TMCH microbiology department for in-house RT-PCR. After in-house RT-PCR, each sample was preserved at −80 °C along with saliva samples.

Reference RT-PCR was performed using a method developed by the National Institute of Infectious Diseases (NIID), Japan, for SARS-CoV-2 [20,21] with purified samples with magLEAD (Precision System Science Co., Ltd., Chiba, Japan). A 200-μL aliquot of each sample was extracted, and 100 μL of purified sample was eluted. For saliva samples, samples were diluted 1:2 with phosphate-buffered saline (PBS) 1x with vortex mixing and then centrifuged for 3 min at 13,000 × *g*, and the supernatant was used as the sample. For RT-PCR, 5 μL of the extracted RNA was used for one-step quantitative RT-PCR with the THUNDERBIRD® Probe One-step qRT-PCR kit (TOYOBO Co., Ltd.) and the LightCycler® 96 Real-time PCR System (Roche Diagnostics KK, Basel, Switzerland). A duplicate analysis for N2 genes was performed for the evaluation of SARS-CoV-2. EDX SARS-CoV-2 Standard (Bio-Rad Laboratories, Inc., Hercules, CA, USA) and sterile purified water (Merck & Co., Inc., Kenilworth, NJ, USA) were used as positive and negative controls, respectively. The calibration curves were generated with 5, 50, and 500 copies/reaction of EDX SARS-CoV-2 Standard.

### Statistical analyses of the rapid antigen tests

The sensitivity and specificity of the antigen tests were calculated with 95% confidence intervals (CIs). The sensitivity stratified by the cycle threshold (Ct) value based on the N2 set of the NIID method was also evaluated. The Ct values according to sample types were compared by Wilcoxon’s signed-rank test.

All statistical analyses were conducted using the R 4.1.2 software program (R Foundation, Vienna, Austria) with the “readxl,” “tidyverse,” and “epiR” packages.

## Results

In total, saliva samples and NP samples were collected from 471 participants during the study period; 455 were from symptomatic participants, and 16 were from asymptomatic participants. For symptomatic participants, the median duration from the symptom onset to sample collection was 1.0 (interquartile range [IQR]: 1.0–2.0) day.

Of the simultaneously obtained saliva samples and NP samples, 140 saliva samples and 143 NP samples were SARS-CoV-2-positive by RT-PCR with the NIID method. Both the saliva and NP samples were positive in 138 participants, whereas saliva samples were positive and NP samples were negative in 2 participants, while saliva samples were negative and NP samples were positive in 5 participants. The median Ct values were 25.5 (IQR: 21.9-28.8) for saliva samples and 17.1 (IQR: 15.5-18.7) for NP samples (p<0.001). The Ct values stratified by the number of positive saliva samples and positive NP samples were 13 (9.3%) for <20, 49 (35.0%) for 20-24, 57 (40.7%) for 25-29, 21 (15.0%) for ≥30 for saliva samples, and 120 (83.9%) for <20, 15 (10.5%) for 20-24, 2 (1.4%) for 25-29, 6 (4.2%) for ≥30 for NP samples. The Ct values of the saliva samples and NP samples are shown in Fig. 2. Antigen testing with saliva samples was performed for all of 471 saliva samples with 3 antigen detection kits. Espline SARS-CoV-2 N required re-tests for 3 samples due to non-reactivities for the positive control line after 20 minutes. There were no other re-tests performed during the study.

**Figure 2.**
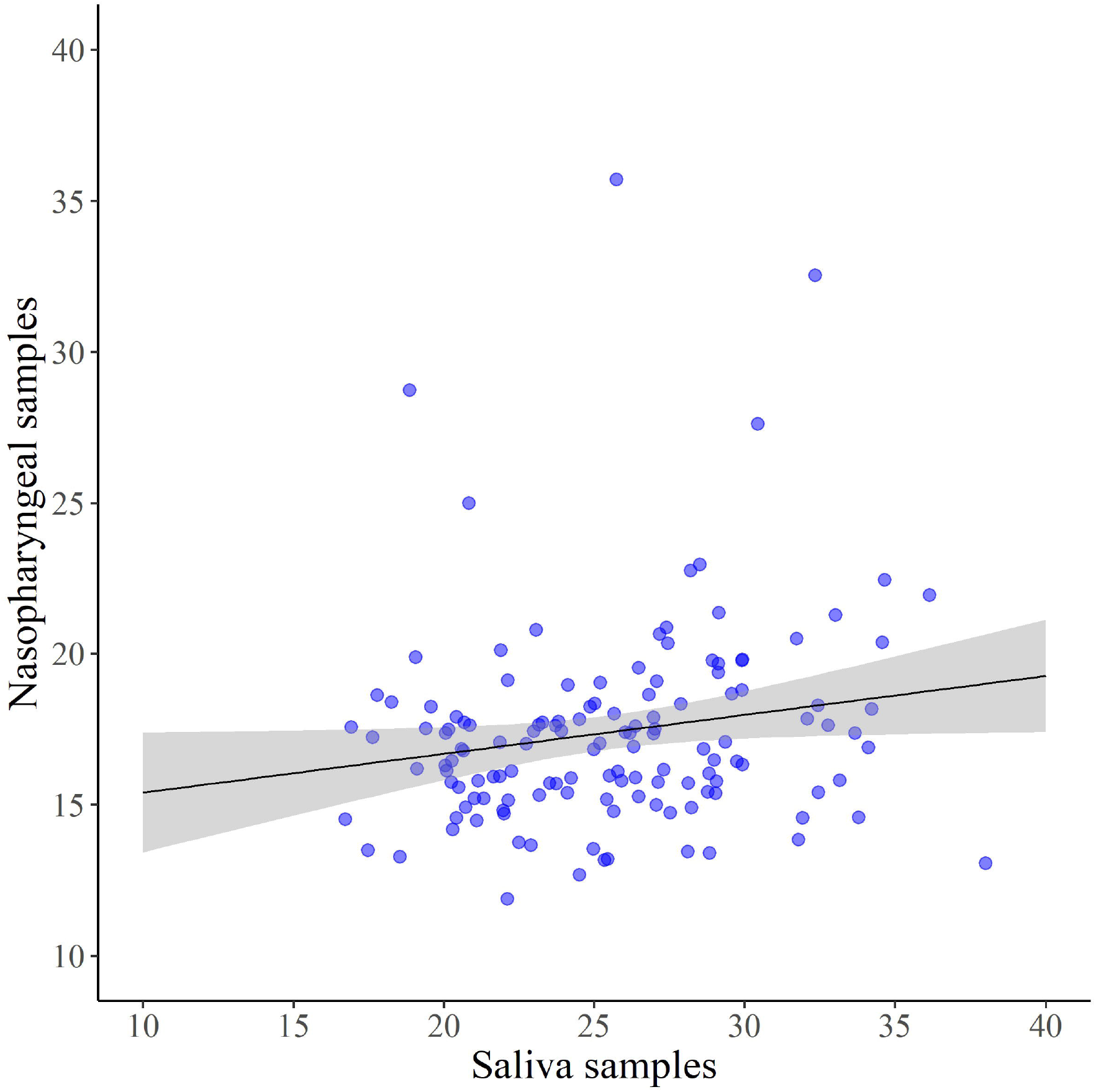
A comparison of cycle threshold values between saliva and nasopharyngeal samples collected from the same participants. A black line with gray area indicates a linear regression line with 95% confidence interval.

Table 1 shows the analytical performance of the three antigen testing kits for the detection of SARS-CoV-2 in saliva when RT-PCR with saliva samples was considered the reference standard. The sensitivity and specificity were 46.4% (95% confidence interval [CI]: 38.0%–55.0%) and 99.7% (95% CI: 98.3%–100%) for ImunoAce SARS-CoV-2 Saliva, 59.3% (95% CI: 50.7%–67.5%) and 99.1% (95% CI: 97.4%–99.8%) for Espline SARS-CoV-2 N, and 61.4% (95% CI: 52.8%–69.5%) and 98.8% (95% CI: 96.9%–99.7%) for QuickChaser Auto SARS-CoV-2, respectively.

**Table 1.** Analytical performance of the three antigen testing kits for the detection of SARS-CoV-2 in saliva (Reference standard: real-time RT-PCR with saliva samples)

| Antigen kits | Total<br>Number | RT-PCR |  | RT-PCR |  | Sensitivity | Specificity |
| --- | --- | --- | --- | --- | --- | --- | --- |
|  |  | (+) | (-) | (+) | (-) |  |  |
|  |  | Ag | Ag | Ag | Ag |  |  |
|  |  | (+) | (-) | (+) | (-) |  |  |
| ImunoAce |  |  |  |  |  | 46.4% | 99.7% |
| SARS-CoV-2 | 471 | 65 | 75 | 1 | 330 | (38.0%-55.0%) | (98.3%-100%) |
| Saliva |  |  |  |  |  |  |  |
| Espline |  |  |  |  |  | 59.3% | 99.1% |
| SARS-CoV-2 N | 471 | 83 | 57 | 3 | 328 | (50.7%-67.5%) | (97.4%-99.8%) |
| QuickChaser Auto |  |  |  |  |  | 61.4% | 98.8% |
| SARS-CoV-2 | 471 | 86 | 54 | 4 | 327 | (52.8%-69.5%) | (96.9%-99.7%) |

RT-PCR, reverse transcription-polymerase chain reaction. Ag, antigen testing.
Data in parentheses are 95% confidence intervals.

Table 2 shows the analytical performance of the three antigen testing kits for the detection of SARS-CoV-2 in saliva when RT-PCR with NP samples was considered the reference standard. The sensitivity and specificity were 44.8% (95% CI:36.4%–53.3%) and 99.4% (95% CI: 97.8%–99.9%) for ImunoAce SARS-CoV-2 Saliva, 57.3% (95% CI: 48.8%–65.6%) and 98.8% (95% CI: 96.9%–99.7%) for Espline SARS-CoV-2 N, and 60.1% (95% CI: 51.6%–68.2%) and 98.8% (95% CI: 96.9%–99.7%) for QuickChaser Auto SARS-CoV-2, respectively.

**Table 2.**
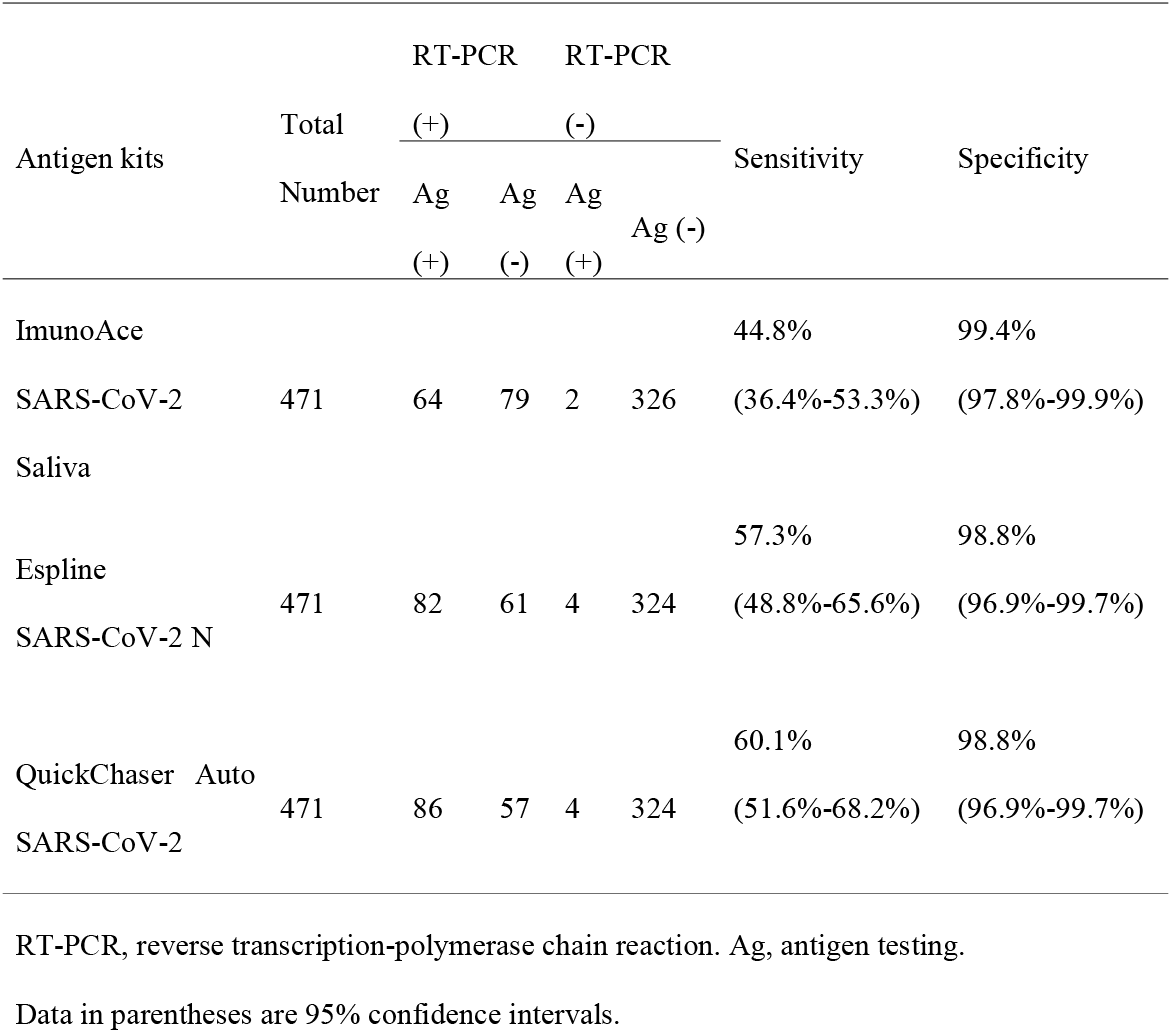
Analytical performance of the three antigen testing kits for the detection of SARS-CoV-2 in saliva (Reference standard: real-time RT-PCR with nasopharyngeal samples)

The sensitivity of the antigen testing kits for the detection of SARS-CoV-2 in saliva stratified by Ct values of saliva samples are shown in Table 3-a. For Ct values of <20, 20–24, 25–29, and ≥30, the sensitivities for ImunoAce SARS-CoV-2 Saliva were 100% (95% CI: 75.3%–100%), 87.8% (95% CI: 75.2%–95.4%), 15.8% (95% CI: 7.5%–27.9%), and 0% (95% CI: 0%–16.1%), respectively; the sensitivities for Espline SARS-CoV-2 N were 100% (95% CI: 75.3%–100%), 93.9% (95% CI: 83.1%–98.7%), 42.1% (95% CI: 29.1%–55.9%), and 0% (95% CI: 0%–16.1%), respectively; and the sensitivities for QuickChaser Auto SARS-CoV-2 were 100% (95% CI: 75.3%–100%), 93.9% (95% CI: 83.1%–98.7%), 47.4% (95% CI: 34.0%–61.0%), and 0% (95% CI: 0%–16.1%), respectively.

**Table 3-a.**
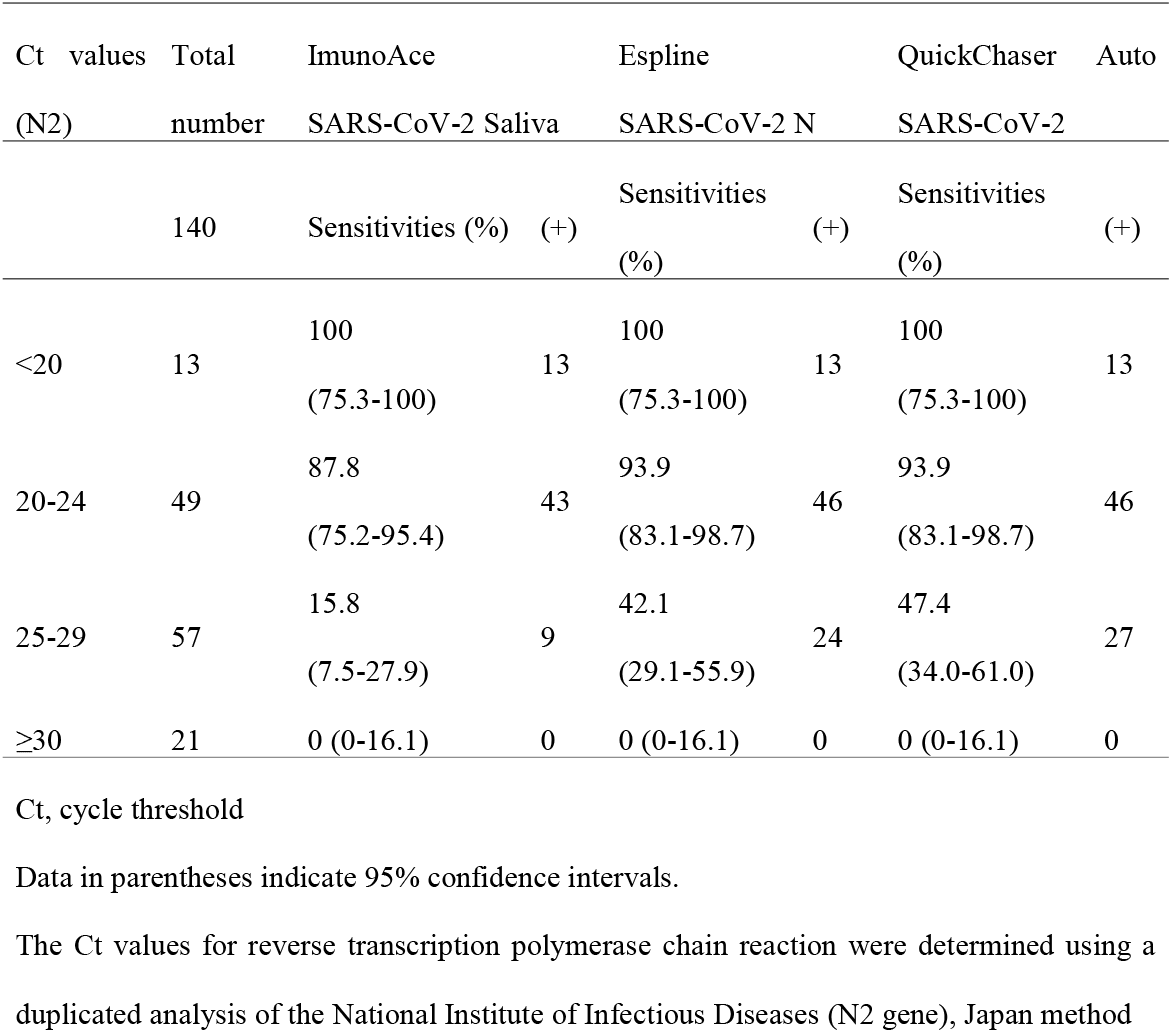
Sensitivities of the antigen testing kits for the detection of SARS-CoV-2 in saliva stratified by Ct values of saliva samples

**Table 3-b.**
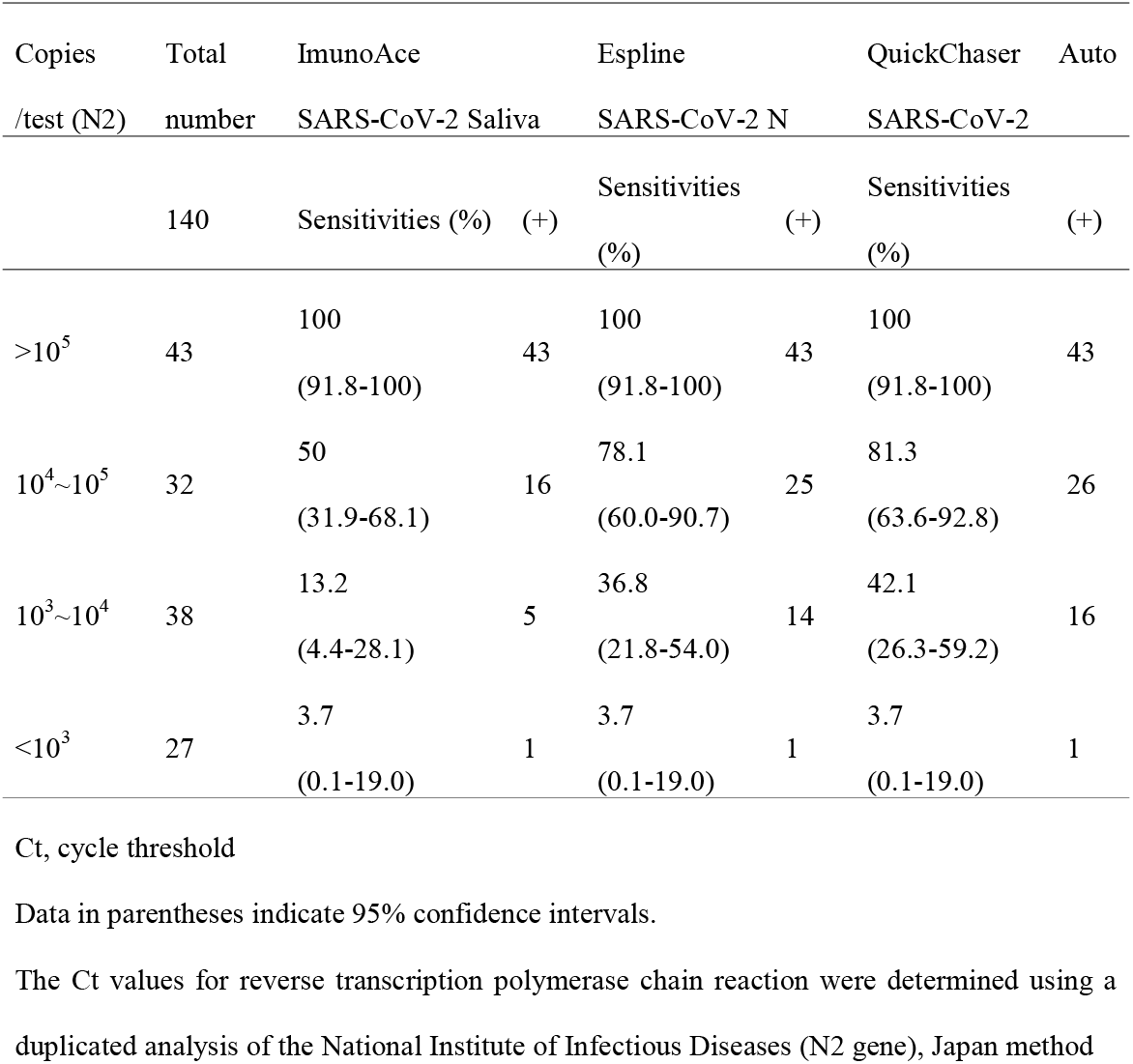
Sensitivities of antigen testing kits for the detection of SARS-CoV-2 in saliva stratified by RNA copies of saliva samples

The sensitivity of the antigen testing kits for the detection of SARS-CoV-2 in saliva stratified by Ct values of NP samples are shown in Table 4-a. For Ct values of <20, 20–24, 25–29, and ≥30, the sensitivities for ImunoAce SARS-CoV-2 Saliva were 50% (95% CI: 40.7%–59.3%), 20% (95% CI: 4.3%–48.1%), 50% (95% CI: 1.3%–98.7%), and 0% (95% CI: 0%–45.9%), respectively; the sensitivities for Espline SARS-CoV-2 N were 64.2% (95% CI: 54.9%–72.7%), 26.7% (95% CI: 7.8%–55.1%), 50% (95% CI: 1.3%–98.7%), and 0% (95% CI: 0%–45.9%), respectively; and the sensitivities for QuickChaser Auto SARS-CoV-2 were 65.8% (95% CI: 56.6%–74.2%), 33.3% (95% CI: 11.8%–61.6%), 50.0% (95% CI: 1.3%–98.7%), and 16.7% (95% CI: 0.4%–64.1%), respectively.

**Table 4-a.**
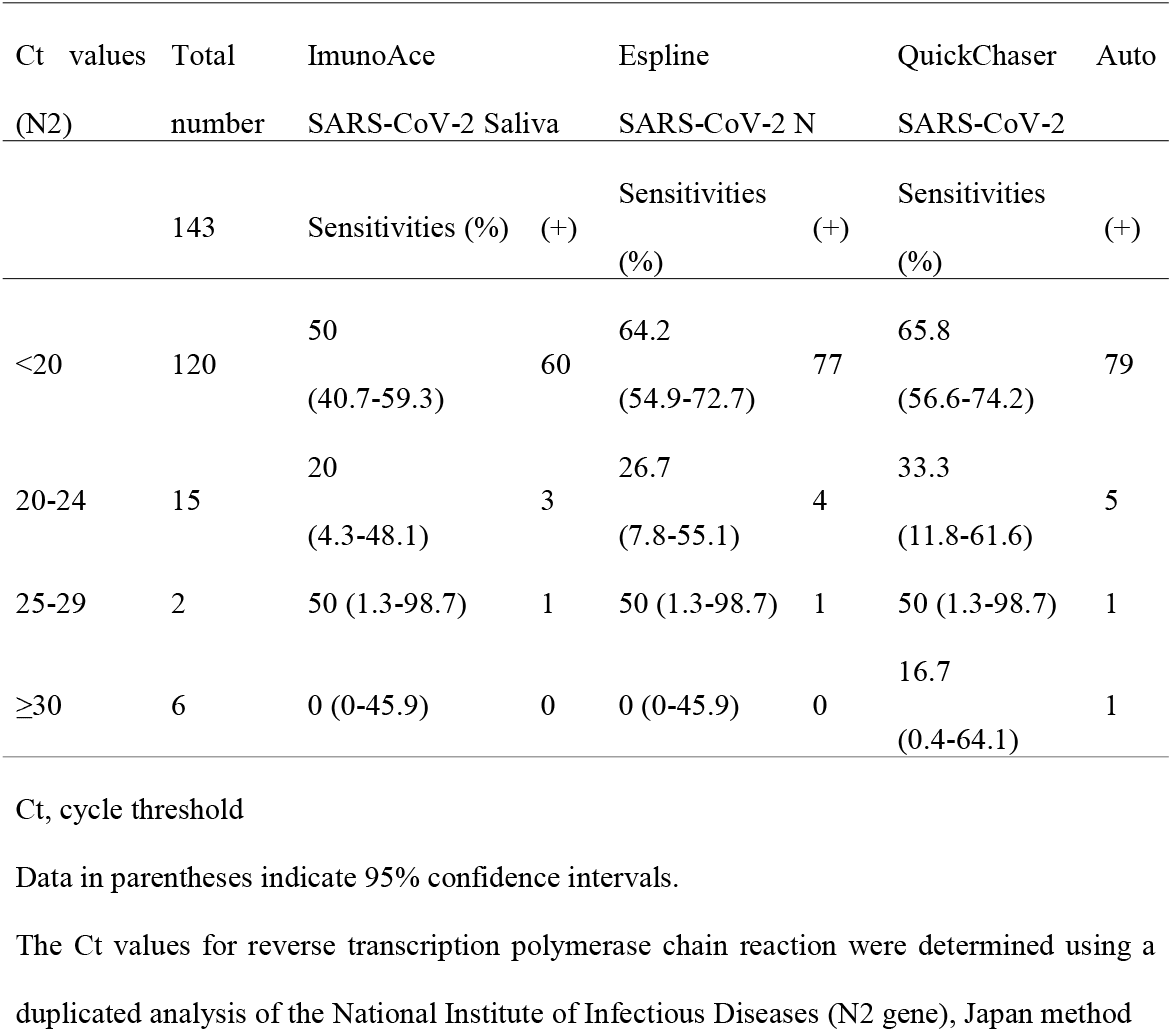
Sensitivities of the antigen testing kits for the detection of SARS-CoV-2 in saliva stratified by Ct values of nasopharyngeal samples

**Table 4-b.**
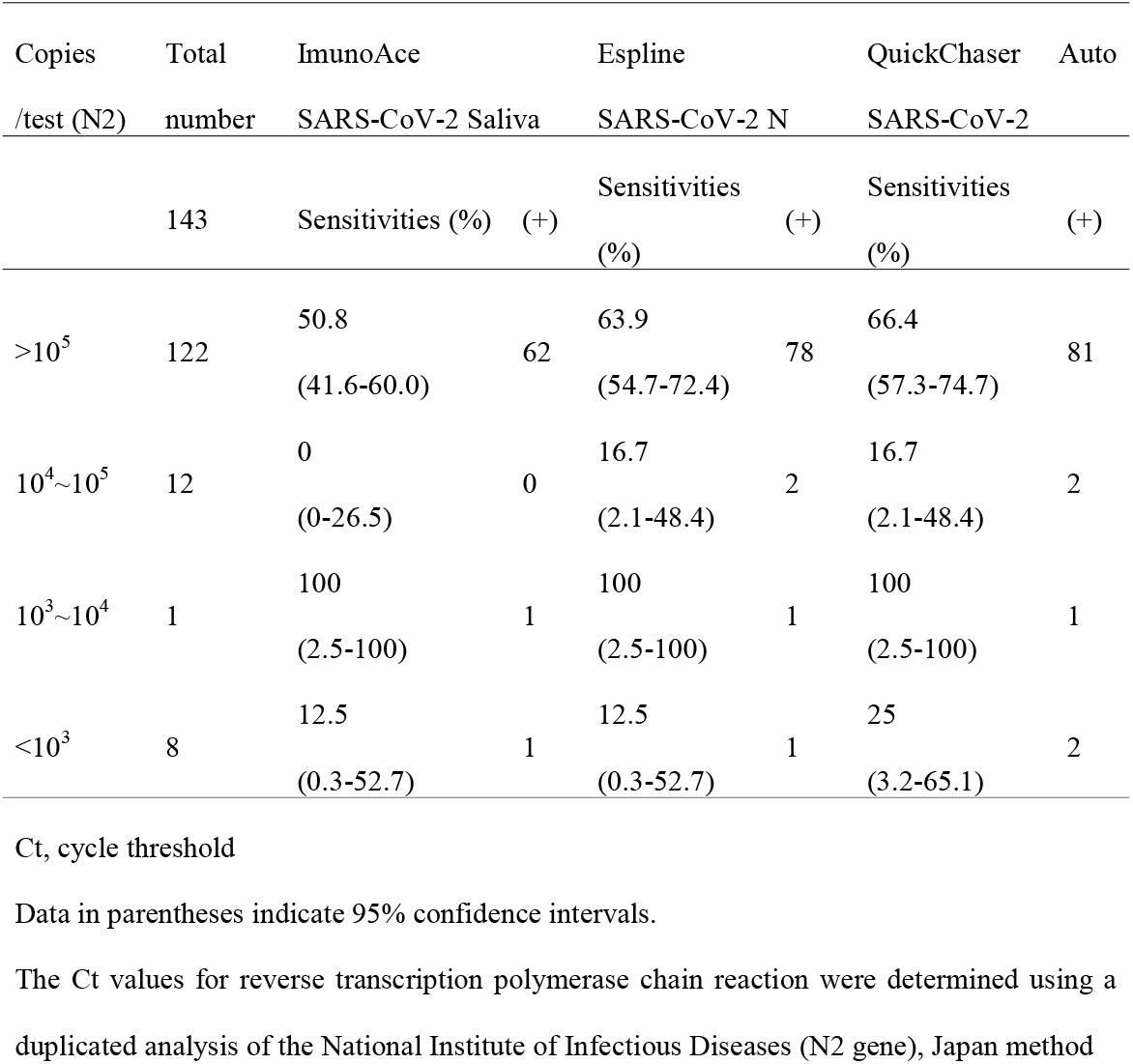
Sensitivities of the antigen testing kits for the detection of SARS-CoV-2 in saliva stratified by RNA copies of nasopharyngeal samples

## Discussion

The current study revealed that each COVID-19 antigen detection kit for saliva had good specificity with infrequent false-positive findings; however, the sensitivities varied among the kits and were much lower than those for NP samples, probably due to the lower viral loads in saliva samples than in NP samples. In this study, high viral loads of SARS-CoV-2 were detected in NP samples in most COVID-19-positive patients, but there were many false-negative results with the antigen detection kits for saliva samples. The sensitivity is >90% for saliva samples with a moderate-to-high viral load (Ct<25), whereas the sensitivity is <70% for high-viral-load nasopharyngeal samples (Ct<20).

While the sensitivities of RT-PCR for the laboratory diagnosis of COVID-19 are similar between NP samples and saliva samples [3,22] including for omicron variants [23], the results of evaluations of saliva qualitative antigen test have not been favorable. Chen et al. performed a systematic review of qualitative antigen tests with saliva in 2022 and found that the pooled sensitivity was 27.4% (95% CI: 8.1%–61.9%), and the pooled specificity was 100% (95% CI: 93.8%–100%) (n=1536), which were significantly lower than those values for quantitative antigen tests (sensitivity: 85.6% [95% CI: 69.2%–94%], specificity 98.9% [95% CI: 94.5%–99.8%]). Yokota et al. evaluated the first-generation Espline kit with 34 frozen positive saliva samples and reported that only 14 samples (41%) were positive [24]. Ishii et al. reported that the sensitivity was 33% (3/9) and the specificity 100% (84/84) [25]. The SD Biosensor saliva antigen rapid test, which is not approved in Japan, was reported to have a better diagnostic performance than other antigen tests in a study of 789 saliva samples. Indeed, Igloi et al. reported that the sensitivity and specificity of the SD Biosensor saliva antigen rapid test were 66.1% and 99.6%, respectively, and the sensitivity increased to 88.6% with a Ct ≤30 cut-off [26].

In the current study, all three evaluated antigen detection kits were newly released and had been adjusted for detection with saliva samples. All of the kits were able to detect SARS-CoV-2 in high-viral-load saliva samples (Ct<25), and Espline SARS-CoV-2 N and QuickChaser Auto SARS-CoV-2 showed better sensitivities in Ct 25–29 samples than ImunoAce SARS-CoV-2 Saliva. According to the Ct-stratified results, the diagnostic performance of Espline SARS-CoV-2 N and QuickChaser Auto SARS-CoV-2 appears similar to that of the SD Biosensor saliva antigen rapid test [26].

For Espline SARS-CoV-2 N, Murakami et al. reported the analytical performance with 60 positive samples and 60 negative samples obtained in 2022, and the sensitivity and specificity were 58.8% (95% CI: 44.2%-72.4%) and 100.0% (95% CI: 94.0%–100.0%). They reported that the sensitivity was 69.8% (95% CI: 53.9%–82.8%) for Ct <30, 92.9% (95% CI: 76.5%–99.1%) for Ct <27, and 100% (95% CI: 80.5%–100%) for Ct <25, which was similar to the current results. The deterioration of sensitivities of qualitative antigen detection kits in saliva samples was considered to be due to the difference in viral loads and sample characteristics between NP samples and saliva samples [23].

Several limitations associated with the present study warrant mention. First, the samples were collected at one site in Japan, and most samples were collected from symptomatic patients soon after the symptom onset. Second, the sample size for asymptomatic individuals was insufficient in this study. Third, most of the positive samples were obtained from participants with high viral loads in the NP, so the sensitivities might be lower at other PCR centers or in asymptomatic individuals.

In conclusion, the current study showed that COVID-19 rapid antigen detection kits with saliva showed high specificities, but the sensitivities varied among kits, and the analytical performance of saliva qualitative antigen detection kits was much worse than that of kits using NP samples.

## Data Availability

All data produced in the present work are contained in the manuscript

## Acknowledgments

We thank Ms. Yoko Ueda, Ms. Mio Matsumoto, Ms. Mika Yaguchi, Ms. Yumiko Tanaka, Ms. Hiromi Tsuruta, Mr. Naoki Tanimura, and the staff of the Department of Clinical Laboratory of Tsukuba Medical Center Hospital for their support in this study. We thank all participating medical institutions for providing their patients’ clinical information.

## References

[1] Hanson KE, Caliendo AM, Arias CA, Hayden MK, Englund JA, Lee MJ, et al. The Infectious Diseases Society of America Guidelines on the Diagnosis of COVID-19: Molecular Diagnostic Testing. Clin Infect Dis 2021;ciab048. https://doi.org/10.1093/cid/ciab557.

[2] takeuchi Y, Akashi Y, Kato D, Kuwahara M, Muramatsu S, Ueda A, et al. Diagnostic performance and characteristics of anterior nasal collection for the SARS-CoV-2 antigen test: a prospective study. Sci Rep 2021;11:10519. https://doi.org/10.1038/s41598-021-90026-8.

[3] Bastos ML, Perlman-Arrow S, Menzies D, Campbell JR. The Sensitivity and Costs of Testing for SARS-CoV-2 Infection With Saliva Versus Nasopharyngeal Swabs: A Systematic Review and Meta-analysis. Ann Intern Med 2021;174:501–10. https://doi.org/10.7326/M20-6569.

[4] World Health Organization. Antigen-detection in the diagnosis of SARS-CoV-2 infection. https://www.who.int/publications/i/item/antigen-detection-in-the-diagnosis-of-sars-cov-2infection-using-rapid-immunoassays. [Accessed 20 October 2022].

[5] Dinnes J, Sharma P, Berhane S, van Wyk SS, Nyaaba N, Domen J, et al. Rapid, point-of-care antigen tests for diagnosis of SARS-CoV-2 infection. Cochrane Database Syst Rev 2022;7:CD013705. https://doi.org/10.1002/14651858.CD013705.pub3.

[6] Safiabadi Tali SH, LeBlanc JJ, Sadiq Z, Oyewunmi OD, Camargo C, Nikpour B, et al. Tools and Techniques for Severe Acute Respiratory Syndrome Coronavirus 2 (SARS-CoV-2)/COVID-19 Detection. Clin Microbiol Rev 2021;34:e00228–20. https://doi.org/10.1128/CMR.00228-20.

[7] Basso D, Aita A, Padoan A, Cosma C, Navaglia F, Moz S, et al. Saliva SARS-CoV-2 antigen rapid detection: A prospective cohort study. Clin Chim Acta 2021;517:54–9. https://doi.org/10.1016/j.cca.2021.02.014.

[8] Akashi Y, Horie M, Kiyotaki J, Takeuchi Y, Togashi K, Adachi Y, et al. Clinical Performance of the cobas Liat SARS-CoV-2 & Influenza A/B Assay in Nasal Samples. Mol Diagn Ther 2022;26:323–31. https://doi.org/10.1007/s40291-022-00580-8.

[9] Akashi Y, Horie M, Takeuchi Y, Togashi K, Adachi Y, Ueda A, et al. A prospective clinical evaluation of the diagnostic accuracy of the SARS-CoV-2 rapid antigen test using anterior nasal samples. J Infect Chemother 2022;28:780–785. https://doi.org/10.1016/j.jiac.2022.02.016

[10] Akashi Y, Kiyasu Y, Takeuchi Y, Kato D, Kuwahara M, Muramatsu S, et al. Evaluation and clinical implications of the time to a positive results of antigen testing for SARS-CoV-2. J Infect Chemother 2022;28:248–51. https://doi.org/10.1016/j.jiac.2021.10.026.

[11] Kiyasu Y, Akashi Y, Sugiyama A, Takeuchi Y, Notake S, Naito A, et al. A Prospective Evaluation of the Analytical Performance of GENECUBE((R)) HQ SARS-CoV-2 and GENECUBE((R)) FLU A/B. Mol Diagn Ther 2021;25:495–504. https://doi.org/10.1007/s40291-021-00535-5.

[12] Kiyasu Y, Owaku M, Akashi Y, Takeuchi Y, Narahara K, Mori S, et al. Clinical evaluation of the rapid nucleic acid amplification point-of-care test (Smart Gene SARS-CoV-2) in the analysis of nasopharyngeal and anterior nasal samples. J Infect Chemother 2022;28:543–547. https://doi.org/10.1016/j.jiac.2021.12.027.

[13] Kiyasu Y, Takeuchi Y, Akashi Y, Kato D, Kuwahara M, Muramatsu S, et al. Prospective analytical performance evaluation of the QuickNavi-COVID19 Ag for asymptomatic individuals. J Infect Chemother. 2021;27:1489–92.

[14] Kurihara Y, Kiyasu Y, Akashi Y, Takeuchi Y, Narahara K, Mori S, et al. The evaluation of a novel digital immunochromatographic assay with silver amplification to detect SARS-CoV-2. J Infect Chemother 2021;27:1493–1497. https://doi.org/10.1016/j.jiac.2021.07.006.

[15] Marty FM, Chen K, Verrill KA. How to Obtain a Nasopharyngeal Swab Specimen. N Engl J Med 2020; 383:e14. https://doi.org/10.1056/NEJMc2015949.

[16] Naito A, Kiyasu Y, Akashi Y, Sugiyama A, Michibuchi M, Takeuchi Y, et al. The evaluation of the utility of the GENECUBE HQ SARS-CoV-2 for anterior nasal samples and saliva samples with a new rapid examination protocol. PLoS One 2021;16:e0262159. https://doi.org/10.1371/journal.pone.0262159.

[17] Suzuki H, Akashi Y, Ueda A, Kiyasu Y, Takeuchi Y, Maehara Y, et al. Diagnostic performance of a novel digital immunoassay (RapidTesta SARS-CoV-2): A prospective observational study with nasopharyngeal samples. J Infect Chemother 2022;28:78–81. https://doi.org/10.1016/j.jiac.2021.10.024.

[18] takeuchi Y, Akashi Y, Kato D, Kuwahara M, Muramatsu S, Ueda A, et al. The evaluation of a newly developed antigen test (QuickNavi-COVID19 Ag) for SARS-CoV-2: A prospective observational study in Japan. J Infect Chemother 2021;27:890–894. https://doi.org/10.1016/j.jiac.2021.02.029.

[19] takeuchi Y, Akashi Y, Kiyasu Y, Terada N, Kurihara Y, Kato D, et al. A prospective evaluation of diagnostic performance of a combo rapid antigen test QuickNavi-Flu+COVID19 Ag. J Infect Chemother 2022;28:840–843. https://doi.org/10.1016/j.jiac.2022.02.027.

[20] Nagura-Ikeda M, Imai K, Tabata S, Miyoshi K, Murahara N, Mizuno T, et al. Clinical Evaluation of Self-Collected Saliva by Quantitative Reverse Transcription-PCR (RT-qPCR), Direct RT-qPCR, Reverse Transcription-Loop-Mediated Isothermal Amplification, and a Rapid Antigen Test To Diagnose COVID-19. J Clin Microbiol 2020;58:e01438–20. https://doi.org/10.1128/JCM.01438-20.

[21] Shirato K, Nao N, Katano H, Takayama I, Saito S, Kato F, et al. Development of Genetic Diagnostic Methods for Detection for Novel Coronavirus 2019(nCoV-2019) in Japan. Jpn J Infect Dis 2020;73:304–307. https://doi.org/10.7883/yoken.JJID.2020.061.

[22] Butler-Laporte G, Lawandi A, Schiller I, Yao M, Dendukuri N, McDonald EG, et al. Comparison of Saliva and Nasopharyngeal Swab Nucleic Acid Amplification Testing for Detection of SARS-CoV-2: A Systematic Review and Meta-analysis. JAMA Intern Med 2021;181:353–360. https://doi.org/10.1001/jamainternmed.2020.8876.

[23] Ursic T, Kogoj R, Sikonja J, Roskaric D, Jevsnik Virant M, Bogovic P, et al. Performance of nasopharyngeal swab and saliva in detecting Delta and Omicron SARS-CoV-2 variants. J Med Virol 2022;94:4704–4711. https://doi.org/10.1002/jmv.27898.

[24] Yokota I, Sakurazawa T, Sugita J, Iwasaki S, Yasuda K, Yamashita N, et al. Performance of Qualitative and Quantitative Antigen Tests for SARS-CoV-2 Using Saliva. Infect Dis Rep 2021;13:742–747. https://doi.org/10.3390/idr13030069.

[25] Ishii T, Sasaki M, Yamada K, Kato D, Osuka H, Aoki K, et al. Immunochromatography and chemiluminescent enzyme immunoassay for COVID-19 diagnosis. J Infect Chemother 2021;27:915–918. https://doi.org/10.1016/j.jiac.2021.02.025.

[26] Igloi Z, Velzing J, Huisman R, Geurtsvankessel C, Comvalius A J IJ, et al. Clinical evaluation of the SD Biosensor SARS-CoV-2 saliva antigen rapid test with symptomatic and asymptomatic, non-hospitalized patients. PLoS One 2021;16:e0260894. https://doi.org/10.1371/journal.pone.0260894.

